# Extraordinary attention, ordinary neglect: the high cost of disaster preparedness and response

**DOI:** 10.1101/2020.10.22.20216275

**Authors:** Robert A. Hahn

## Abstract

**Background:** Funds allocated to disaster preparedness and response in the U.S. have grown rapidly in recent decades. This analysis examines the ratio of cost per outcome of public health events classified as disasters and those not classified as disasters, e.g., smoking-related morbidity and mortality.

**Methods:** Mortality is taken as an outcome metric; the validity of this measure is assessed by examination of ratios of tangible and intangible costs of disaster and non-disaster outcomes to mortality from two conditions, using available data. The relative allocation of CDC funding to disaster and non-disaster events is assumed to conservatively represent the U.S. overall relative funding allocation.

**Results:** Non-disaster deaths are 2,500 more likely than disaster deaths; we allocate 370 times more funding per disaster death than we do per non-disaster death.

**Conclusion:** The rationality of this implicit decision be reconsidered.

## Introduction

Disasters happen. Some are deliberately caused by humans, others perhaps unintentional consequences of human action, some apparently natural. As the boy scouts sagely advise, preparedness can be helpful—allowing the mitigation of harm and the seizure of opportunity. Here I ask what happens to non-disasters, i.e., health events that are not disasters, while we are preparing for and responding to disasters. How do we decide the how to allocate resources between these two classes of events? During disasters, resources from normal public health activities may be redirected to disaster response.[1] What do we forgo when we respond to these crises; what are the opportunity costs? In this exploration, I focus on disaster efforts in the U.S., but similar questions can be raised globally. I exclude the COVID-19 pandemic from this analysis because neither costs nor consequences are yet clear; moreover, available preparedness appears to have been largely ignored.

While terminology is not used consistently among agencies, disaster analysts generally distinguish several phases in events of disaster, including preparedness, response, and recovery.(REF)

The National Disaster Recovery Framework 2016 (https://www.fema.gov/media-library-data/1466014998123-4bec8550930f774269e0c5968b120ba2/National_Disaster_Recovery_Frame-work2nd.pdf) includes the following definitions:

**“Prevention:** The capabilities necessary to avoid, prevent, or stop a threatened or actual act of terrorism. Within national **preparedness**, the term “prevention” refers to preventing imminent threats.

**Response:** The capabilities necessary to save lives, protect property and the environment, and meet basic human needs after an incident has occurred.

**Recovery:** The capabilities necessary to assist communities affected by an incident to recover effectively.”

While we focus here on preparedness and response, the issue of recovery is raised in addressing question 3 below. We ask five questions and use rough indices with available data to answer them.

1. What distinguishes disaster from non-disaster?
2. What is the relative health burden of disaster and non-disaster events in the U.S.
3. Is mortality a reasonable index for the relative burden of disaster and non-disaster events?
4. What is the relative allocation of resources for disaster and non-disaster events in the U.S. What are the ratios of resources/disaster and /non-disaster events in the U.S., and how do these two ratios compare, i.e., how does the allocation of resources for disaster outcomes compare with that for non-disaster outcomes—a ratio of ratios?
5. It is difficult to ascertain the cost of averting a death associated with disaster, but I ask what is the cost of averting deaths from several non-disaster causes of deaths?

## Methods

Mortality is used as an estimate of the relative burden of disaster and non-disaster events, following categories from the standard ICD coding. To validate the use of mortality as an overall outcome metric, reports on the relative tangible and intangible costs of disaster and non-disaster events are compared with mortality rates from the same events. The relative allocation of spending on disaster and non-disaster events in the budget of the U.S. Centers for Disease Control and Prevention is assessed as an indication of relative health cost allocation in the U.S. The ratios of CDC budget allocations to U.S. mortality for disaster and non-disaster events are compared. The cost of reducing mortality from specific causes of death are estimated from available studies.

## Findings

### 1. Distinguishing disaster from non-disaster

Public health disasters (or emergencies) are events that are unexpected (at least by their victims), rapid in onset, large in consequences (at least locally), and, thus, beyond the coping capacity of usual resources.[2-6] Though the terminology is not used consistently, some agencies distinguish “emergencies,” which demand resources beyond routine practice, but which can be addressed with available resources, from “disasters,” which result in greater damage and demand more resources than locally available.[5] For purposes of this analysis, we will group both in the more severe category, i.e., disasters. Some events are disasters because they threaten contagion and have major impact. What also clearly count as disasters are: major natural events—large area fires, droughts, hurricanes, floods, tsunamis, earthquakes; terrorist attacks, war, and genocide; famine and large scale epidemics. Some of what we have thought of as natural disasters are increasingly recognized to have roots in human activity, with unintended and sometimes intended consequences. In the U.S., both can be given federal support by Presidential declaration using the Stafford Disaster Relief and Emergency Assistance Act.(https://www.doi.gov/sites/doi.gov/files/uploads/Stafford_Act_pdf.pdf)

### 2. The relative health burden of disaster and non-disaster in the U.S

To estimate the relative public health burden of events that are disasters and not disasters, we use mortality data because they are available, reliable, and a major measure of health. Mortality is an indicator of additional suffering (in the form of morbidity) and can serve as rough estimator of broader health phenomena. The assumption here is not that the focus of preparedness and prevention is mortality, or that other outcomes, e.g., morbidity or the destruction of property are comparatively less important, but only that mortality is a reasonable proxy for all outcomes combined. We test this assumption below.

Among all 2.74 million deaths in the U.S. in 2016—the latest data available, less than 0.01% were attributed to terrorism, and less than that each for natural disasters and conflict; say that disaster-associated mortality accounts for 0.04% of total US mortality (https://ourworldinda-ta.org/is-it-fair-to-compare-terrorism-and-disaster-with-other-causes-of-death). In most years since 1900 there were fewer than 500 deaths from natural disasters in the U.S.(ref) Perhaps additional disastrous deaths are attributed to other causes, but even doubling or tripling the known proportion, the mortality attributable to disaster in the U.S. in 2016 is relatively small. To look at the ratio of disaster to non disaster events in another way, the likelihood of dying of a non disaster event in the U.S. is approximately 2,500 times (0.9996/0.0004) that of dying of a disaster event. Cardiovascular diseases and cancers combined—the two leading causes—account for 57% of U.S. deaths in that year (https://ourworldindata.org/is-it-fair-to-compare-terrorism-and-disaster-with-other-causes-of-death). The top ten causes combined account for 88.7% of all deaths.

The more than 480,000 deaths annually caused by tobacco, mostly consumed voluntarily, are not regarded as disaster—1,315 deaths each day and 17.6% of US mortality. But imagine that no smoking-related deaths occurred for 51 weeks in the year, then 9,205 occurred (at the rate of 1,315 deaths daily) during a single week. In this case, about 2% of smoking-related deaths might provoke a rapid investigation and disaster response, while 100% of smoking-related deaths spread throughout the year does not. In 2016, more that 20,000 Americans were killed in motor vehicle crashes, more than half of them were not wearing seat-belts. There were 38,600 firearms deaths in the U.S.[7] These are not classified as disasters.

### 3. Is mortality a reasonable index of the relative burden of disaster and non-disaster events?

We use mortality as a metric to compare the relative costs of disaster and non-disaster preparedness and prevention. While mortality statistics in the U.S. are comprehensive and generally accurate sources of health events, they may not equally represent the burdens of disaster and non-disaster events. Clearly, the costs associated with smoking and with natural disasters differ fundamentally—the costs of smoking are those associated with the living smoker and the additional costs of natural disaster accrue predominantly after disaster-related deaths. The issue of comparison, however, is whether the burdens of both are close enough in magnitude, whenever they occur, for the deaths to constitute a reasonable metric for comparison of costs of prevention and preparedness for the two event types.

To explore this issue and assess the validity of mortality as an index of burden, we compare forms of burden for smoking-related deaths [8] and the consequences of natural disasters [9] in a high income nation for which such data are available—Australia, where intangible well as tangible outcomes associated with smoking and with natural disasters have been assesses Direct tangible costs are “those incurred as a result of the hazard event and have a market value such as damage to properties, infrastructure, vehicles and crops…. Indirect tangible costs … arise from the consequences of the damage and destruction such as business disruption, clean-up emergency relief and recovery costs, and network disruptions…. Intangible costs arise from loss of life and from pain and suffering.”[9] Intangible costs may include family violence, mental health, alcohol and drug misuse, unemployment, educational outcomes, and injury and loss of life. These costs are difficult to assess, but efforts are made. Mental health, for example, may be assigned costs by assessing treatment costs and effects on work loss. We can also measure changes in mental health or other morbidity before, during, and after disaster events. Clearly, these approaches do not include the suffering involved.

One report [9] estimates the tangible and intangible costs of two major natural disasters in Australia—the 2010–2011 Queensland, Australia floods and the 2009 Black Saturday bushfires in Victoria state which (combined) resulted in 209 deaths and 414 injuries, an estimated tangible loss of $9.8 billion and an intangible loss of $11.3 billion; the mortality component of this amount is $3.6 billion. The other report [8] estimates the tangible and intangible costs of cigarette smoking in Victoria in 1998/99 (noting similar costs for Australia as a whole; Table 1). In Victoria, 4,747 deaths are attributed to smoking in 1998/99, tangible losses associated with smoking are estimated to have been $1,593.6 billion and intangible losses $3,456.3 billion—an estimate of mortality costs alone. In both disaster and non-disaster conditions, intangible costs are substantially greater than tangible costs. For purposes of the present analysis, what matters is the ratio comparing two other ratios—tangible and intangible costs/death for disaster and non-disaster events. The ratio of disaster to non-disaster mortality costs/death are 0.14 tangible costs, 0.023 for intangible costs, and 0.060 for tangible and intangible mortality costs combined. Assuming that this comparison is representative of the difference between disaster and non-disaster events, the higher relative costs associated with (non-disaster) smoking-related mortality suggests that our estimates of the relative costs of disaster and non-disaster preparedness and pre vention are conservative—by approximately 16 fold.

**Table 1.** Comparing Tangible and Intangible Consequences of Disaster and Non-DIsaster.

| Event (Source) | Smoking in Victoria/Australia, 98/99 (Collins 2006) | Natural disaster Australia—2015 Queensland floods and Black Saturday Fires (Deloitte 2016) |
| --- | --- | --- |
| Death | 4,747 | 209 |
| Injury/victims |  | 414 |
| Mental health | | 86,200 persons/ lifetime cost \$1 billion |
| Tangible | \$1,593.6 Billion | \$9.8 billion |
| Intangible—mortality | \$3,456.3 Billion | \$3.6 billion |
| —morbidity | NA | \$7.7 billion |
| Tangible costs/death (x1 b) | 0.336 | 0.047 |
| Intangible mortality costs/death (x1 b) | 0.728 | 0.017 |
| (Tang + Intang)/deaths (x1 b) | 1.064 | 0.064 |

### 4. What is the relative allocation of resources for disaster and non-disaster events in the U.S., and how do these ratios compare?

To examine the proportion of resources the federal government spends on disaster and non-disaster conditions, we examine the budget of the CDC—the major public health agency in the U.S. There are other resources spent on both disasters and non-disasters in the U.S.; but we assume the CDC proportions are indicative of the allocation of federal resources for disaster and non-disaster events in the U.S. A society’s allocation of economic resources is thought to reflect its values.[10]

The 2019 budget for the CDC was $10.921 billion. “Public health preparedness and response” were allocated $1.402 billion (12.8%); so, approximately $9,519 billion remained for non-disaster spending (https://www.hhs.gov/sites/default/files/fy-2019-budget-in-brief.pdf). This means that per death that occurred in 2016, $1,272,700 was spent for preparedness per disaster death and $3,500 was spent for preparedness per nondisaster death. Almost 370 times the preparedness funding was allocated for each disaster death (i.e., ((1.402/0.0004)/(9.519/0.9996)) as for each non disaster death. Some disasters, e.g., the Ebola outbreak of 2015-17 and the Zika outbreak of more recent years are provided additional funding beyond the allotted CDC budget, so that the ratio is still greater. If the U.S. military budget and the budgets of FEMA and the Department of Homeland Security are regarded as components of disaster preparedness and the avoidance of deliberate mass harm to our population, the ratio becomes astronomical.

Posid and colleagues [1] have estimated the proportion of CDC personnel that is diverted from routine public health work during a declared public health emergency. This estimate does not include the personnel routinely monitoring events to determine the onset of an emergency, and additional resources may also be called from other agencies. In the events that Posid examined, approximately 9.5% of CDC personnel are called for a mean of 118 days per event. The proportion of CDC person-time allocated to the average event can thus be estimated to be 0.095 × 118/365 = 3% per emergency event. The CDC reports 55 full emergency responses over a 10 year period, 2003 - 12,[11] thus approximately 5.5 per year, in which case, on average, approximately 16.5% (3% x 5.5) of CDC personnel would have been occupied in these activities. Approximately 3/4th of these were for events in the U.S. Man-made disaster accounted for 16.4% of the activations, including 5.5% classified as bioterrorism. This does not count “109 other occasions, the EMP [Emergency Management Program] was used to support emergency responses that did not require full EOC [the Emergency Operations Center] activation, and the EMP also conducted 30 exercises and drills” at additional cost. Nor does it count the background activity maintaining emergency assessment and surveillance activities. For example, “During 2004– 2012, EOC watch officers triaged an average of 23,303 requests per year (range: 14,633– 38,812),”—more than 60 per day.[11]

### 5. What is the cost of averting deaths from several non-disaster causes?

It may be that the small number of disaster deaths in the U.S. is in part a consequence of preparedness. Intense attention to a few cases of Ebola in 2014 in the U.S. most likely averted great harm. Because of their military strategic importance, it is difficult to find statistics on the number of deaths and other harms averted by preparedness activities in the U.S. The cost of these preventions is beyond public access, in reasonable part to obscure prevention work. Some available data may give a lower bound estimate on number of events averted. It is reported that 38 terrorist events in stages of planning or execution have been averted since 9/11, approximately two per year (Wikipedia: https://en.wikipedia.org/wiki/List_of_unsuccessful_terrorist_plots_in_the_United_States_post-9/11). The number of terrorist events deterred by our preparedness, including our surveillance apparatus, is impossible to estimate. Similarly, we may also expect that the number of non-disaster incidents and deaths—tobacco-related deaths and motor vehicle injuries would be larger than it is were it not for effective prevention programs and policies supported by expenditures on these matters.

It is also quite plausible that it costs more to prevent a disaster-associated death than to prevent non-disaster related death. Disasters are often unpredictable or at least unpredicted, and those that are intentional are often designed to be both unpredicted and impactful. Because much of the activity and cost of disaster prevention is hidden from public scrutiny, it is not possible to estimate the cost of the extensive prevention activity per event prevented. Estimation is feasible for non-disaster events. We know the cost of NOT getting a smoker to quit, not wearing a seatbelt; these costs are high. For example, the societal costs of cigarette smoking in the U.S. is approximately $300 billion annually, which includes direct medical costs as well as lost productivity to smokers themselves and to those exposed to second-hand smoke; per adult smoker, that is more than $7,000 annually. (CDC: https://www.cdc.gov/tobacco/data_statistics/fact_sheets/economics/ econ_facts/index.htm#anchor_1548357936093)

What is the cost of converting a smoker into a non-smoker? Asaria et al.[12] have estimated the cost in several low- and middle-income nations (thus not including the U.S.) of several anti-smoking interventions with demonstrated effectiveness in reducing rates of smoking-related mortality. The average cost of three anti-smoking programs combined (i.e., increased tobacco taxes, smoke-free workplaces, and requirements for labelling of tobacco products, public awareness campaigns, and a ban on tobacco advertising, promotion, and sponsorship) is an average of $0.26 per capita population per year for 10 years, i.e., $2.60 total, for a 15% reduction in the prevalence of smoking. Let us say that the costs of such programs per capita in the U.S. are double the highest annual cost in the Asaria study, i.e., $0.72 in Poland. The prevalence of smoking among adults in the U.S. is currently approximately 15.1%.[13] Thus, the cost of these programs per smoker (rather than per capita) is approximately $95.36 (($0.72 × 2 × 10)/0.155). And, if the prevalence of smoking is reduced by 15%, the cost per smoker who ceases to smoke is $636.00 ($95.36/0.15). Spending $636 to convert a smoker into a non smoker, thus saving $7,000 per year is an exceptional bargain.

## Discussion

This analysis rests on many assumptions, some of which may be questioned. One is that what are reported as disaster-related deaths, and thus, by subtraction, counted as non-disaster deaths, are actually representative of what are counted as disasters and non-disasters. We have also assumed that the CDC budget is representative of the relative expenditures for disaster and non-disaster preparedness/prevention in the U.S. What counts as disaster preparedness is a matter that may involve the military, whose budget is orders of magnitude larger than that of public health and perhaps not fully transparent; addition of this factor would only increase the issue of imbalance raised here. Another assumption, addressed above is whether mortality is a good metric by which to measure the burdens of disaster and non-disaster events. Data to address the relative cost for disaster and non-disaster events are hard to come by; sources are chosen because they are available, plausible, and valid, not to present a particular outcome.

Findings from psychological studies of decision-making may partially explain the extensive differential allocation of resources. Humans routinely overestimate the magnitude and frequency of rare events and underestimate the magnitude and frequency of common events,[14] a phenomenon also referred to as “probability neglect”[15] and “overreaction to fearsome events.”[15] Rare and threatening events receive disproportionate attention in the media which distorts their relative importance. We may allocate resources accordingly. Experimental efforts to correct misjudgments of relative magnitude are ineffective. These fears may be costly.

## Conclusion

The findings from this analysis are rough, but even if inaccurate by an order of magnitude, they indicate a questionable alignment. We are approximately 2,500 times more likely to die of a non-disaster event than of a disaster. Yet we spend roughly 370 times as much to prevent disaster-related events as we do to prevent non-disaster-related events. What the relative allocation of resources to disaster versus non-disaster events should look like is not clear. I suspect that the current massive gap is problematic and may not correspond with shared values. We may be driven by irrational fears to spend inordinate attention to the (rare) extraordinary to the relative neglect of the (common) ordinary. I do not know if there is a rational way to decide appropriate allocation, but I believe it is worthwhile to attempt to make the comparison, to be aware of the gap, and to consider this question. We can ask if the ratio we estimate here is one we find reasonable and acceptable. Data and analysis should play prominent roles in the allocation of societal resources.

## Data Availability

available from author

